# Reduced antibody acquisition with increasing age following vaccination with BNT162b2: results from a large study performed in the general population aged 12 to 92 years

**DOI:** 10.1101/2022.05.18.22275209

**Authors:** Lotus L. van den Hoogen, Mardi C. Boer, Abigail Postema, Lia de Rond, Mary-lène de Zeeuw-Brouwer, Inge Pronk, Alienke J. Wijmenga-Monsuur, Elske Bijvank, Caitlyn Kruiper, Lisa Beckers, Marjan Bogaard-van Maurik, Ilse Zutt, Jeffrey van Vliet, Rianne van Bergen, Marjan Kuijer, Gaby Smits, W. M. Monique Verschuren, H. Susan J. Picavet, Fiona R.M. van der Klis, Gerco den Hartog, Robert S. van Binnendijk, Anne-Marie Buisman

## Abstract

Vaccine-induced protection of the population against severe COVID-19, hospitalization and death is of utmost importance, especially in the elderly. However, limited data are available on humoral immune responses following COVID-19 vaccination in the general population across a broad age range. We performed an integrated analysis of the effect of age, sex and prior SARS-CoV-2 infection on Spike S1-specific (S1) IgG concentrations up to three months post BNT162b2 vaccination. 1·735 persons, eligible for COVID-19 vaccination through the national program, were recruited from the general population (12 to 92 years old). Sixty percent were female and the median vaccination interval was 35 days (interquartile range, IQR: 35-35). All participants had seroconverted to S1 one month after two doses of vaccine. S1 IgG was higher in participants with a history of SARS-CoV-2 infection (median: 4·535 BAU/ml, IQR: 2·341-7·205) compared to infection-naïve persons (1·842 BAU/ml, 1·019-3·116) after two doses, p<0.001. In infection-naïve persons, linear mixed effects regression showed a strong negative association between age and S1 IgG one month after the first vaccination (p<0.001) across the entire age range. The association was still present after the second vaccination, but less pronounced. Females had higher S1 IgG than males after both the first and second vaccination (p<0.001); although this difference was lower after the second dose. In persons with an infection history, age nor sex was associated with peak S1 IgG. As IgG decreased with age and time since vaccination, older persons may become at risk of infection, especially with escape variants such as Omicron.

## Introduction

Understanding and monitoring immune responses following COVID-19 vaccination is essential to protect the population against severe COVID-19. The mRNA BNT162b2 vaccine was the first COVID-19 vaccine to be approved by the FDA and EMA, and is currently used in 159 countries around the world [1]. Generally, the vaccination strategy for BNT162b2 consists of a primary series of two vaccinations 3-6 weeks apart. Early studies on BNT162b2 demonstrated high vaccine efficacy against COVID-19 infection after completing the full vaccination schedule [2]. This has subsequently been confirmed by real-world data, considering protection against severe COVID-19 [3, 4].

The elderly population is especially at risk for severe COVID-19, hospitalization and death. Age-related changes of the immune system, referred to as immunosenescence, contribute to the increased susceptibility to infectious diseases and reduced efficacy of vaccination in elderly persons [5, 6]. Others have shown impaired immune responses following COVID-19 vaccination when comparing data from elderly nursing home residents to those from younger healthcare workers [7, 8]. However, apart from difference in age range, these specific groups differ considerably from the general population in terms of health status. Further data for elderly people outside of nursing home settings are lacking [9]. Thus, data on the peak antibody concentrations post vaccination in the general population as a whole remains limited [10, 11]. This information however is essential for further COVID-19 vaccination strategies to be able to better protect vulnerable groups, like the elderly, against severe disease. In addition, immunity of the general population as a whole tempers virus transmission which further lowers the overall disease burden.

In the Netherlands, COVID-19 vaccinations were offered from early 2021 onwards. Initially, priority was given to frontline healthcare workers and individuals at high risk for severe COVID-19 due to long-term health conditions. Afterwards, the general population was invited for vaccination, descending by age. Currently, everyone 5 years or older has access to free COVID-19 vaccination. The most commonly used vaccine for all ages is BNT162b2, except those aged 60-64. This group mostly received Vaxzevria as their primary vaccination series due to national policies at the time.

In this article we discuss COVID-19 serological findings across 1·735 persons in the Dutch general population with an age range of 12 to 92 years following a primary series of BNT162b2. We determined the effect of age and sex on antibody acquisition up to one month following one and two doses of BNT162b2, and stratified results by SARS-CoV-2 infection history. Subsequently, the decay in antibody concentrations from one to three months following two doses of BNT162b2 vaccination were analysed for persons aged 50 and over.

## Methods

### Study population

Data from two observational, longitudinal COVID-19 vaccination cohort studies with the same study design were combined: one focusing on adolescents and adults (12 to 60 years old) and one on the ageing population (50+ years old). Participants were included in the study if they planned to receive COVID-19 vaccination or completed the primary vaccination series within the last 28 days, as this was the primary endpoint of the study. See Supplementary Material for further details on study design, recruitment and inclusion and exclusion criteria.

### Sample collection

Finger-prick blood samples and questionnaires were taken at four time points; prior to COVID-19 vaccination (Pre-vacc), 28 days after the first vaccination (Dose 1), 28 days after the second vaccination (Dose 2), and three months after the second vaccination (Month 3). Month 3 data was only available for the 50+ cohort. Questionnaires covered demographic factors, COVID-19 vaccination information (brand and dates), and SARS-CoV-2 testing information (if applicable). Finger-prick blood samples were self-collected in microtubes and returned by mail. Once received at the laboratory at the National Institute of Public Health and the Environment (RIVM), the Netherlands, serum was isolated from each sample by centrifuge and stored at -20°C until sample processing.

### Immunoglobulin G detection

Total immunoglobulin G (IgG) antibody concentrations to Spike S1 and Nucleoprotein were measured simultaneously using a bead-based assay as previously described [12]. IgG concentrations were expressed as binding antibody units per mL (BAU/ml) using 5-parameter logistic interpolation of the International Standard for human anti-SARS-CoV-2 immunoglobulin (20/136 NIBSC standard) [13]. The threshold for seropositivity was set at 10.1 BAU/ml for Spike S1 [14] and 14.3 BAU/ml for Nucleoprotein [15].

### Statistical analyses

All statistical analyses were performed in RStudio (version 4.1.3) [16]. Age in years at first vaccination was used. Persons who received one or two doses of BNT162b2 were selected. Persons who received BNT162b2 in a heterologous schedule were excluded from analysis. Samples were included if they were within -7 or +7 days from day 28 (Dose 1), -14 or +14 days from day 28 (Dose 2), and -14 or +14 days from three months (Month 3). Persons with serological evidence of a SARS-CoV-2 infection history (i.e., seropositive to Spike S1 at Pre-vacc) and/or a self-reported positive SARS-CoV-2 test performed by local health authorities prior to their vaccination schedule were analysed separately. SARS-CoV-2 confirmatory testing was available free of charge to anyone with COVID-19 related symptoms or those who were in close contact with someone who had tested positive according to national guidelines in the Netherlands. Persons who reported a positive SARS-CoV-2 test during – or following, for the 50+ cohort – their vaccination schedule were excluded. Participants who were seronegative for Spike S1 but seropositive for Nucleoprotein at Pre-vacc were also excluded as their infection-status was considered non-conclusive. Flowcharts of the number of participants and available samples are shown in Supplementary Figure 1.

The Mann-Whitney-test was used to compare antibody concentrations between groups. A linear mixed effects regression model with a random intercept per participant was used to determine the effect of timepoint, age and sex on S1 IgG concentrations using *lme4* (version 1.1.28 [17]). A model with these explanatory variables as well as an interaction term between timepoint and age and timepoint and sex was created. Separate models were built for infection-naïve persons and those with a history of SARS-CoV-2 infection. The Pre-vacc timepoint was not included in the model for infection-naïve participants as we assumed no effect of age and sex on seronegative IgG S1 concentrations. Backward stepwise model selection was performed and the model with the lowest Akaike Information Criterion (AIC) was selected using the step function within *lmerTest*. S1 IgG concentrations were log10-transformed for regression models. Two additional models were built to assess antibody production following first vaccination, and then to assess decay following completion of the primary vaccination series. The former model used data from infection-naïve participants with measurements at both Dose 1 and Dose 2, and the latter model used data from infection-naïve participants with measurements at both Dose 2 and Month 3. In addition to age and sex, we explored the effect of antibody concentrations produced at a previous timepoint on the fold-change to the next timepoint; e.g. the effect of IgG S1 concentrations at Dose 1 on the fold-change of antibody concentrations between Dose 1 and Dose 2. Fold-change was calculated with IgG S1 concentrations on the linear scale. Linear regression was conducted using age, sex and IgG concentration as explanatory variables, as well as the interactions between these variables. Final model selection was based on the AIC, as described above.

### Ethics statement

Ethical approval was obtained through The Medical Research Ethics Committee Utrecht for both the 50+ population cohort (NL74843.041.21, EudraCT: 2021-001976-40), and for the adolescent and adult cohort (12-60 years) (NL76440.041.21, EudraCT: 2021-001357-31). All participants provided written informed consent. In the case of participants aged 12 till 16, both parents also provided written informed consent.

## Results

### Study population

A total of 1·735 BNT162b2-vaccinated participants were included and 4·925 measurements were available: 1·377 at Pre-vacc, 1·429 at Dose 1, 1·425 at Dose 2 and 694 at Month 3 (50+ cohort only). As vaccines were rolled-out per age group from old to young according to the national vaccination campaign, older persons were less likely to have a pre-vaccination measurement. Twenty-two participants dropped out of the study after enrolment. Overall, more females than males were included (1·036; 60% vs 687; 40%). The median interval between the two vaccination doses was 35 days (interquartile range, IQR: 35-35) and did not differ between age groups (Supplementary Table 1). In the ages 20-59 years old, approximately 20% had a SARS-CoV-2 infection prior to vaccination, followed by 12% for the ages 12-19, 5% for the ages 60-79 and 0% in 80+ (Supplementary Table 1).

### Higher Spike S1-specific IgG in persons with a history of SARS-CoV-2 infection

At Dose 1, 97% (1·204/1·235) were seropositive among infection-naïve persons and 100% (193/194) among those with an infection prior to vaccination. Seropositivity at Dose 1 decreased with age (Supplementary Table 2). Seropositivity was 100% at Dose 2 irrespective of age and prior infection status. For all ages, median S1 IgG in infection-naïve increased from 0 BAU/ml Pre-vacc (IQR: 0-1), to 146 BAU/ml at Dose 1 (72-290; Pre-vacc vs. Dose 1: p<0.001), and further to 1·842 BAU/ml at Dose 2 (1·019-3·116; Dose 1 vs. Dose 2: p<0.001), Figure 1. Persons who experienced a SARS-CoV-2 infection prior to vaccination increased from 73 BAU/ml Pre-vacc (29-147), to 3·293 BAU/ml at Dose 1 (1·191-5·751; Pre-vacc vs. Dose 1: p<0.001) and 4·535 BAU/ml at Dose 2 (2·341-7·205; Dose 1 vs. Dose 2: p=0.002); their S1 IgG was higher compared to infection-naïve participants at each timepoint (p<0.001).

**Figure 1:**
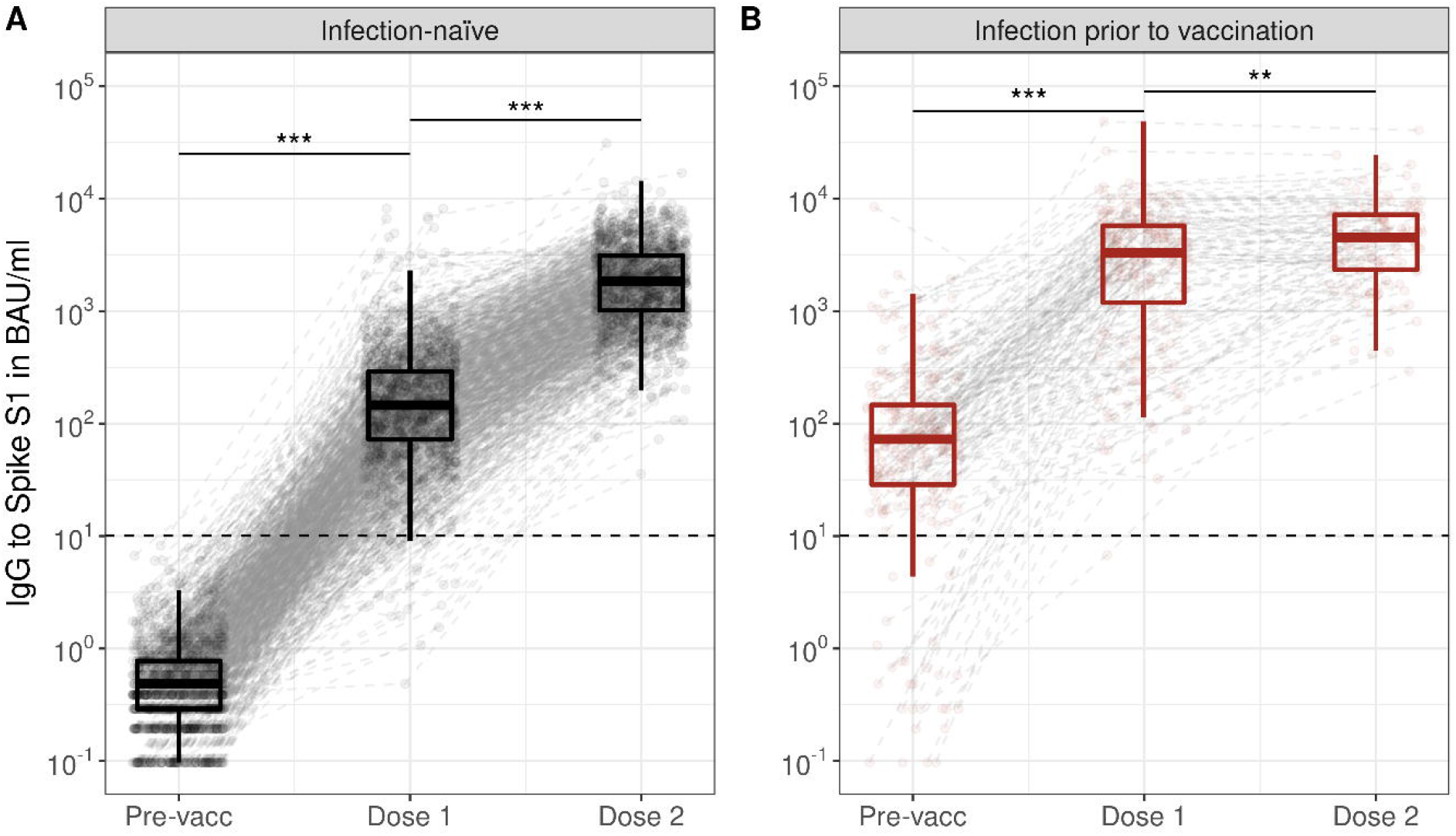
Spike S1-specific IgG kinetics following the primary series of BNT162b2 vaccination in infection-naïve participants (A) and participants with a SARS-CoV-2 infection prior to vaccination (B). Boxplots show results for all participants at each timepoint, while dots and dashed grey lines show measurements and their trajectory between timepoints per participant. Results are shown for total of 1·500 infection-naïve participants and 235 participants with a SARS-CoV-2 infection history; measurements were not available at every timepoint for each participant. Spike S1 IgG concentrations were expressed in international binding antibody units (BAU) using the 20/136 NIBSC standard and were taken prior to vaccination (Pre-vacc), one month following the first (Dose 1) and one month following the second vaccination dose (Dose 2). The horizontal dashed line represents the threshold for seropositivity to Spike S1. Asterisks indicate p-values from Wilxocon-Mann Whitney test (unpaired), when the Wilcoxon signed ranks test was performed in only paired samples all p-values were <0.001. IgG: immunoglobulin G; BAU: binding antibody units; ***: p<0.001; **: p=0.002.

### Age is a strong indicator of Spike S1-specific IgG acquisition following vaccination

BNT162b2-induced S1 IgG upon vaccination decreased in concentration with age, with the highest concentrations in the age group 12-19 years old (Dose 1: median 526 BAU/ml, 282-770 and Dose 2: median 4·198 BAU/ml, 2·692-6·131). We observed stepwise drops per age decade to a median concentration of 45 BAU/ml (16-113) for Dose 1 and a median concentration of 672 BAU/ml (366-1·304) for Dose 2 in the 80-92 age group (see Figure 2 and Supplementary Table 2). There was a strong negative association between age and IgG concentration at Dose 1 and Dose 2 (Table 1 and Figure 3). The strength of the negative association between age and S1 IgG concentration decreased between Dose 1 and Dose 2, indicating greater antibody acquisition with increasing age between Dose 1 and Dose 2 (coefficient of interaction term age*Dose 2: 0.004; p<0.001), Table 1. Females had higher IgG S1 concentrations compared to males at Dose 1 (0.093; p<0.001), but this difference was smaller at Dose 2 (coefficient of interaction term Female*Dose 2: -0.044; p=0.048). Lower concentrations of S1 IgG at Dose 1 resulted in greater fold-change between the first and second vaccination dose (Supplementary Table 3 and Supplementary Figure 2). In persons with an infection history, age nor sex were associated with peak S1 IgG concentrations (Figure 2, Supplementary Figure 3, Supplementary Table 4).

**Figure 2:**
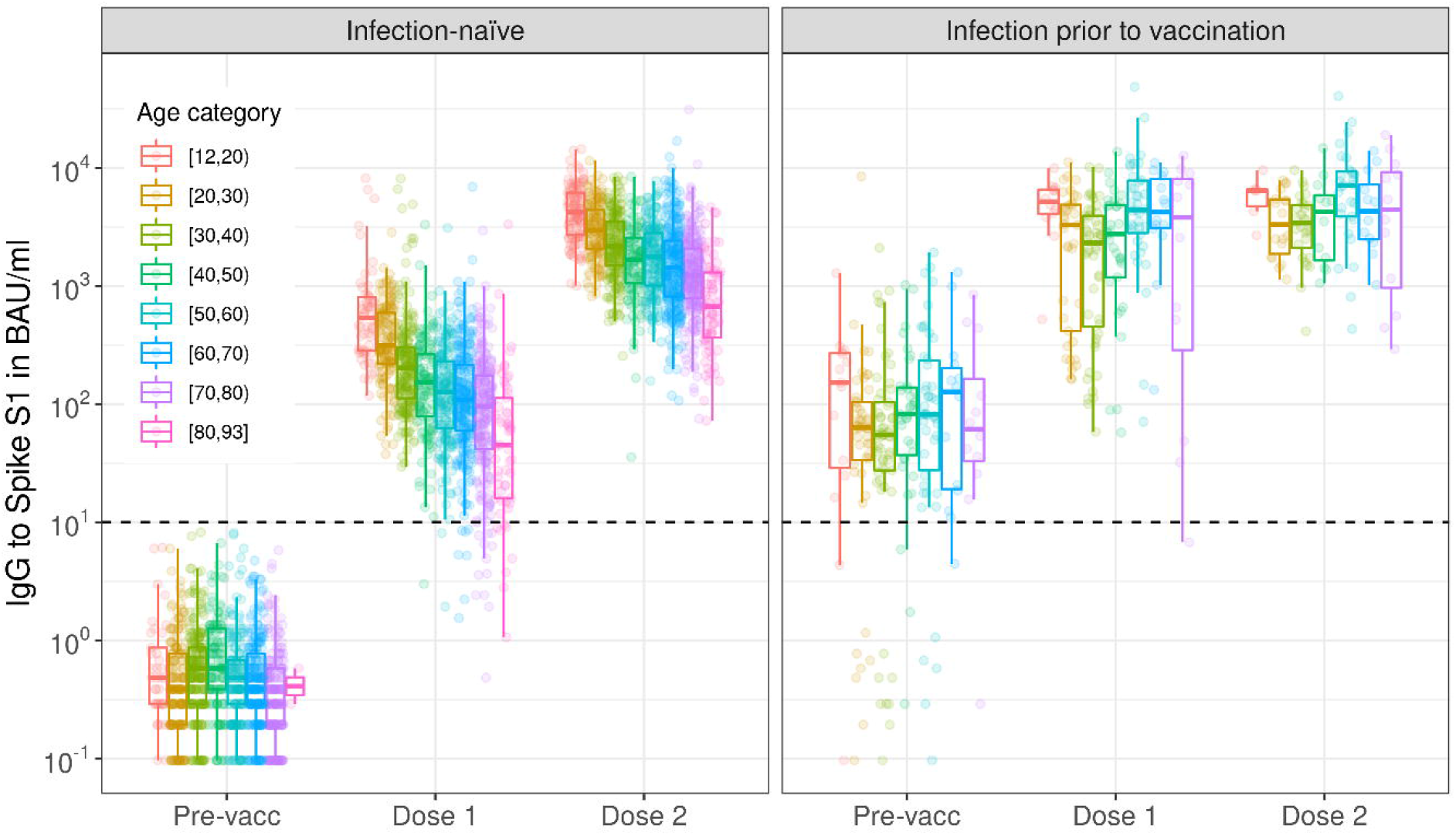
Spike S1-specific IgG kinetics by age category following the primary series of BNT162b2 vaccination in infection-naïve participants (A) and participants with a SARS-CoV-2 infection prior to vaccination (B). Boxplots show results by age group at each timepoint, while dots show individual measurements. Results are shown for total of 1·500 infection-naïve participants and 235 participants with a SARS-CoV-2 infection history; measurements were not available at every timepoint for each participant (see Supplementary Table 2). Spike S1 IgG concentrations were expressed in international binding antibody units (BAU) using the 20/136 NIBSC standard and were taken prior to vaccination (Pre-vacc), one month following the first (Dose 1) and one month following the second vaccination dose (Dose 2). The horizontal dashed line represents the threshold for seropositivity to Spike S1. IgG: immunoglobulin G; BAU: binding antibody units.

**Figure 3:**
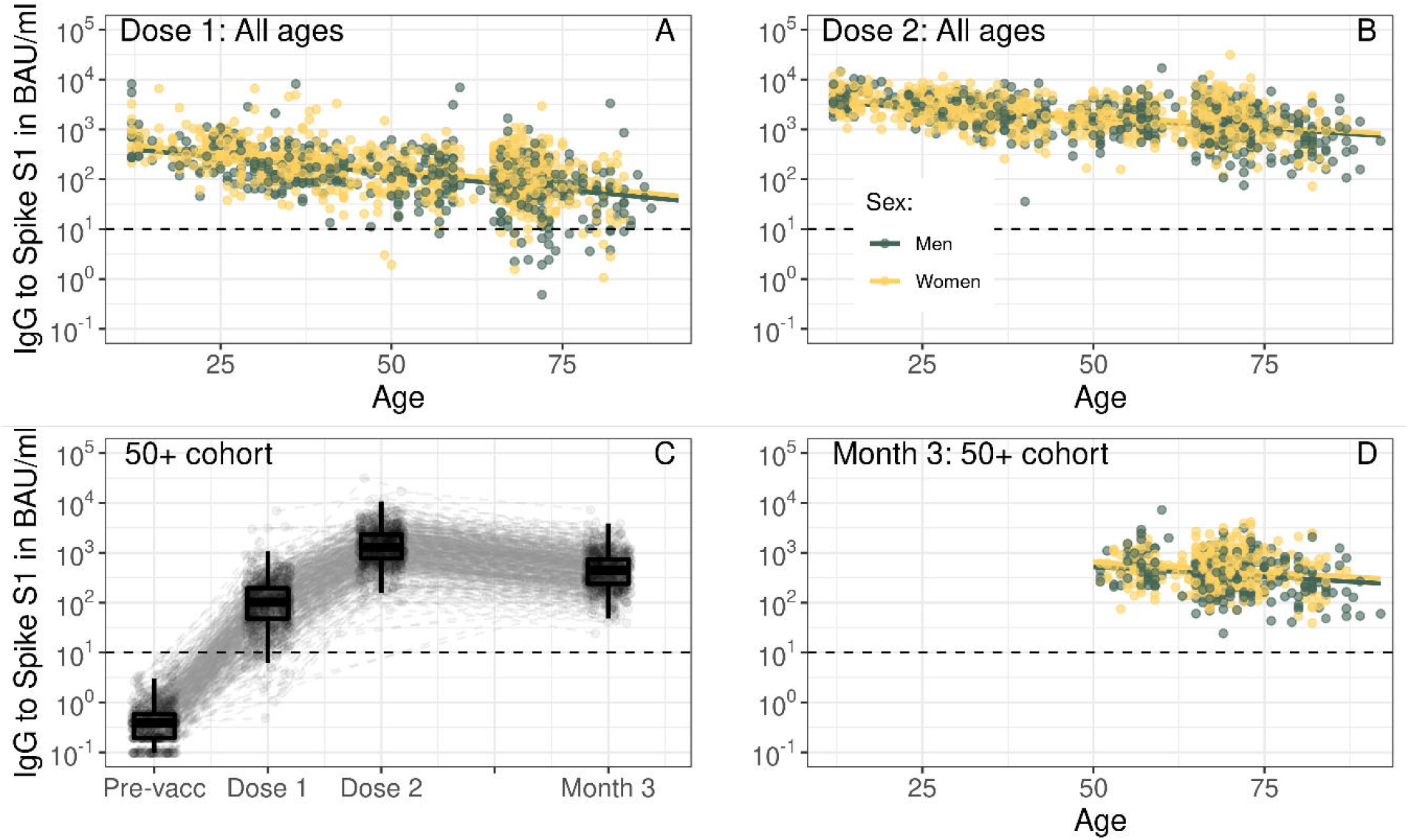
Spike S1-specific IgG by age in years per timepoint (A,B,D) and kinetics following the primary series of BNT162b2 vaccination in infection-naïve participants up to three months following the second vaccination dose (C). In (A, B, D) fitted lines represent the linear association between IgG concentration and age from linear mixed effects regression results (see Table 1 and Supplementary Table 4), while dots represent individual measurements. Results are shown separately for males (green) and females (yellow). In (C) boxplots show results for all participants at each timepoint, while dots and dashed grey lines show measurements and their trajectory between timepoints per participant. In (A-D) Spike S1 IgG concentrations were expressed in international binding antibody units (BAU) using the 20/136 NIBSC standard and were taken prior to vaccination (Pre-vacc), one month following the first (Dose 1), one month following the second (Dose 2) or three months following the second vaccination dose (Month 3). In (A-B) results are shown for total of 1·448 unique infection-naïve participants across all ages with S1 IgG measurements available at Dose 1 and/or Dose 2, while in (C) results are shown for 749 unique infection-naïve participants in the 50+ cohort and in (D) results are shown for 725 unique infection-naïve participants in the 50+ cohort with measurements available at Dose 2 and/or Month 3. The horizontal dashed line represents the threshold for seropositivity to Spike S1. IgG: immunoglobulin G; BAU: binding antibody units.

**Table 1:** Linear mixed effects regression results for Spike S1 IgG concentrations up to one month following two doses of BNT162b2 for infection-naïve persons. Model results for 1·448 unique participants with S1 IgG measurements available at one month after the first (Dose 1) or second vaccination dose (Dose 2). Three persons reporting their sex as “other” were excluded from the model.

|  | Coefficient | 95% CI | p-value |
| --- | --- | --- | --- |
| **Timepoint** |  |  |  |
| - Dose 1 | Ref. |  |  |
| - Dose 2 | 0.891 | 0.822,0.960 | <0.001 |
| **Age in years** | -0.013 | -0.014,-0.012 | <0.001 |
| **Sex** |  |  |  |
| - Male | Ref. |  |  |
| - Female | 0.093 | 0.049,0.136 | <0.001 |
| **Age\*Dose 2** | 0.004 | 0.003,0.006 | <0.001 |
| **Sex\*Dose 2** |  |  |  |
| - Male*Dose 2 | Ref. |  |  |
| - Female*Dose 2 | -0.044 | -0.087,-0.000 | 0.049 |

### Spike S1-specific IgG decay is reduced with increasing age in persons 50 years and over

For infection-naïve persons in the 50+ cohort, median S1 IgG decreased from 1·304 (772-2·318) at Dose 2 to 440 (239-736) at Month 3 but all participants were still seropositive at Month 3 (Figure 3C and Supplementary Table 2). The strength of the negative association between age and S1 IgG was lower at Month 3 compared to Dose 2, indicating slower antibody decay with increasing age between Dose 2 and Month 3 (coefficient for interaction term age*Month 3: 0.004; p<0.001, Figure 3D and Supplementary Table 5). The coefficient for higher S1 IgG in females compared to males remained the same between Dose 2 and Month 3 (0.102; p<0.001). For antibody loss – i.e., the fold-change between one and three months following the second vaccination – higher concentrations of IgG S1 at Dose 2 resulted in a greater loss (Figure 4 and Supplementary Table 6). For females, antibody loss was faster than for males at high concentrations of Dose 2 IgG S1 but slower at low concentrations of Dose 2 IgG S1. In persons with an infection history, neither age nor sex were associated with S1 IgG concentrations at both post-vaccination timepoints (Supplementary Figure 3 and Supplementary Table 5).

**Figure 4:**
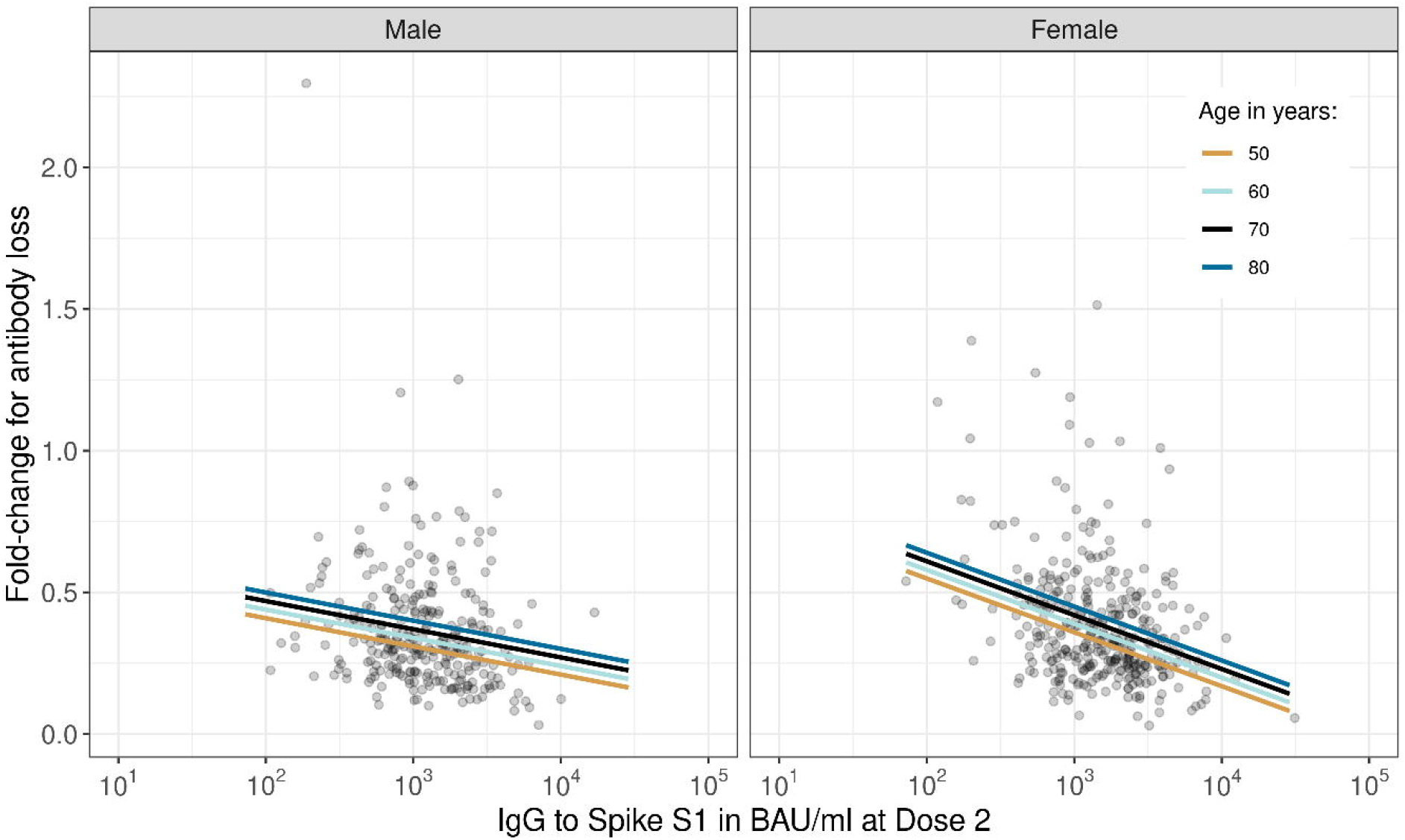
The fold-change in Spike S1-specific IgG to represent antibody loss between one and three months following the second dose of BNT162b2 by IgG S1 concentration at one month following the second dose. Fitted lines represent the linear association between fold-change and age from linear mixed effects regression results for four ages (see Supplementary Table 6), while dots represent individual measurements. Results are shown separately for males and females. This only includes participants who participated at both timepoints (i.e., one month and three months following the second dose of BNT162b2); n=633.

## Discussion

Here we show high peak IgG responses to Spike S1 following two doses of BNT162b2 in infection-naïve persons from the general population across a wide age range of 12 to 92 years. At the time of writing, this study is unique as it includes people of all ages, enabling direct comparison of S1 antibodies in adolescents, adults and the elderly. The large numbers of participants in this study allowed us to demonstrate a strong negative association between IgG S1 concentrations and age after the first dose of BNT162b2. This association was less pronounced but still present after the second dose. In general, the primary vaccination series resulted in high peak antibody concentration even in the oldest persons.

We showed that IgG concentrations one month after the first dose were negatively associated with age. This may be due to an overall superior B cell function in younger persons, possibly involving a broader repertoire of naïve B cells. These cells are more quickly activated upon exposure to the novel antigen introduced by the vaccine. Additionally, antigen uptake, processing, presentation, and signalling of innate cells, as well as recruiting T cell help are more efficient in the young. This may contribute to higher antibody concentrations upon initial vaccination in younger persons [18, 19]. However, the increase in antibody concentrations induced by the second vaccine dose was greater with increasing age, partly compensating the lower antibody acquisition after the first dose. One explanation might be that high concentrations of circulating IgG – seen in younger persons – could partially blunt the response to the second vaccine dose and thus limit restimulation of antibody production following the second vaccine dose. Specifically, considering the relatively short vaccination interval of five weeks. Another explanation might be that peak antibody production was not yet achieved in older persons one month after the first vaccination dose. Interestingly, we also demonstrated that antibody loss between one and three months following the second dose was slower with older age in the 50+ cohort. This could be explained by IgG half-life which affects lower IgG values in older persons less than higher IgG values in younger persons, and by differences in underlying short-lived plasma cell responses and memory B cell formation across age ranges [19, 20].

Other studies have previously shown the inverse relationship between age and antibody concentrations following one dose of BNT162b2 in the infection-naïve general population or healthcare workers [7, 11]. However, observations of antibody concentrations following two doses are contradictory, with some studies reporting limited to no effect [7, 9] and others strong associations with age [10, 21-24]. The lack of an observed age-association after two doses reported in some studies might be explained by the serological assay used, the time window of sampling since the second dose, the time interval between the two doses, and/or the sample size of the study. The sample size in this study was large across all ages with well-defined sampling times post-vaccination. However, it should be noted that persons of 60-64 years old were underrepresented in our study due to national guidelines to vaccinate with Vaxzevria. The pronounced age-effect demonstrated in this study was still present one month after the second dose, and for the 50+ cohort at three months following the second dose. This is in line with results from a recent pre-print showing lower vaccine effectiveness against SARS-CoV-2 infection to the Delta and Omicron BA.1 variants with increasing age in the Netherlands [25]. Nevertheless, infection risk is determined by a complex interaction of infection pressure, behaviour, and the immune system’s response to invasion of the virus, which of course constitutes more than antibody concentrations alone.

To date, most data on antibody responses following BNT162b2 vaccination in the elderly (80+) were derived from nursing home residents, the most frail group of older individuals [7, 8]. The ageing population, however, shows a huge heterogeneity in health status, with early signs of ageing already occurring in the 5^th^ decade (40-50) of life [26]. The diversity between individuals is caused by a complex interplay of physiological and maybe pathological changes in metabolism, organ function, the innate and adaptive immune system, and differences in exposure to risk factors [18]. In nursing home residents aged 80 and over the seroconversion rate for Spike S1 following the primary series of BNT162b2 was 89% [8]. Here, we showed 100% seropositivity in the elderly general population, outside of nursing home settings, up to three months following the second vaccine dose. Reassuringly, this implies that in a large part of the older general population immune response is better when compared to older persons in nursing homes. Similarly, Parry et al. also studied persons ages 80 and over in the general population and showed a seroconversion rate of 96% after two doses of BNT162b2 (n= 100) [9].

As previously described by others, we showed that females had slightly higher antibody concentrations than males following BNT162b2 vaccination [10, 11, 22]. This has been hypothesized to be due to sex steroid hormones or other genetic factors [5, 6]. Also in line with previous findings, we showed higher antibody concentrations in participants with a history of SARS-CoV-2 infection [10, 11, 27]. This finding has led to a national recommendation in The Netherlands of requiring a single dose to complete the primary vaccine series in persons with previous SARS-CoV-2 infection. We found that peak IgG S1 concentrations one month after the second dose were not affected by age or sex in persons with an infection history. However, in the 50+ cohort at three months following the second dose there was more variation in IgG concentrations, but also fewer observations available across a smaller age range. Still, the persistence of antibodies may differ for those receiving one or two doses following infection, or with age and sex.

In conclusion, we showed a strong negative association between BNT162b2-vaccination induced S1 IgG concentration and age. High concentrations of circulating antibodies are important for neutralizing SARS-CoV-2 infection, as also recently exemplified by escape variants such as Omicron. Monitoring the persistence or decay of immune responses following vaccination is pivotal, specifically to evaluate antibody concentrations associated with immune protection in the elderly population. Such knowledge could support vaccination strategies to sustain optimal population immunity. Therefore, future work will focus on vaccine-induced SARS-CoV-2 humoral and cellular responses over time across age, infection history, number of received doses and health status.

## Data Availability

All data produced in the present study are available upon reasonable request to the authors

## Acknowledgements

We would like to thank all participants.

