## Supplementary Material for "Reduced antibody acquisition with increasing age following vaccination with BNT162b2: results from a large study performed in the general population aged 12 to 92 years"

***Contents***

*Cohort studies: study design, recruitment, and inclusion and exclusion criteria………………………. page 2*

*Supplementary Figures…………………………………………………………………………………………………………… page 5*

*Supplementary Tables……………………………………………………………………………………………………………. page 11*

***Cohort studies: study design, recruitment, and inclusion and exclusion criteria***

Data from two COVID-19 vaccination cohort studies were combined. Details on the study design and population are described in detail below. Participants were included in the study if they planned to receive COVID-19 vaccination and were willing to participate. Those who had already received vaccination were also eligible if final recruitment occurred before the end of the 28 day period following the second dose. Exclusion criteria are detailed below.

*Adolescent and adult cohort*

Adolescents 12 to 17 years of age were invited to participate by simple random sampling from the National Population Register (Basis Registratie Personen; BRP). Forty-thousand adolescents were invited and the target was 150 participants. Adolescents were vaccinated with Comirnaty according to Dutch national guidelines. Participants were excluded if they participated in a vaccination or medication clinical trial and/or were severely immunocompromised (see Supplementary Table).

Adults 18 to 60 years of age were invited to participate through multiple channels:

- By simple random sampling from the BRP (n=40,000)
- Via participation in other studies by the Dutch National Institute of Public Health and the Environment (RIVM). Participants who consented to be contacted for additional research were invited.
- By spontaneous applications from interested citizens.

The initial target was to include 150 participants per age group (18-29, 30-44 and 45-60) and vaccine type (Comirnaty, Vaxzevria, Janssen or Spikevax). However, Comirnaty became the most widely used vaccine in the Netherlands and thus no restrictions to enrolment were applied after inviting potential participants. Participants were excluded if they participated in a vaccination or medication clinical trial and/or were severely immunocompromised (Supplementary Table).

*50+ cohort*

The Doetinchem Cohort Study (DCS) is an ongoing prospective study that began in 1987 in Doetinchem, eastern Netherlands [1, 2]. The first study round in 1987 included a sex-stratified random sample of 20-59 years old inhabitants of the city of Doetinchem. Adults 50-90 years of age who had participated in round 6 (2013-2017) of DCS and consented to be contacted for additional research were invited to participate (n=3,147).

**Supplementary Table: Overview of study population, inclusion and exclusion criteria as well as period of enrolment per cohort.**

|  | **Adolescent and adult cohort**  **(18-60 years)** | **50+ cohort**  **(50-92 years)** |
| --- | --- | --- |
| Invited population | Random sample of national population register (n=80,000)  Participants in other studies by the Dutch National Institute of Public Health and the Environment (n~9,500)  Spontaneous enrollment via website | Participated in round 6 of Doetinchem Cohort Study [1, 2] (n=3,147) |
| Inclusion criteria | Planning to receive/received COVID-19 vaccination*  Able and willing to participate and sign IC | Planning to receive/received COVID-19 vaccination*  Able and willing to participate and sign IC |
| Exclusion criteria | Participation in a phase I/II/III preregistration vaccination trial or a phase I/II/III medicine (pre-registration) trial  Belonging to a high risk group for COVID-19 already studied in a risk group vaccination study that this study provides as a comparison for **  Any other immune deficiency through disease  Active or past immunosuppressive or immune modulating medication***  Women who are pregnant or breastfeeding  Having (functional) asplenia  Receipt of blood products or immunoglobulin, within 3 months of study entry  Receipt of organ transplant not mentioned in high risk group list | None |
| Period of enrolment | April to October 2021 | March to August 2021 |

*Latest enrolment: within one month following second vaccination.

**Primary (inherited) immune deficiency, severely decreased kidney function (defined as Chronic Kidney Disease stage 4 or 5; eGFR<30), treatment by dialysis or recipient of a kidney transplant, pulmonary disease for which the patient will receive or has received a lung transplant, autoimmune disease (e.g. MS, rheumatoid arthritis, IBD, SLE etc.), Down syndrome, (known) infection with Human Immunodeficiency Virus (HIV), cancer patients and patients with active cancer treatment (including hormone therapy), receipt of chemotherapy in the last 3 years and/or any history of cancer immune therapy, haematological patients, such as haematological malignancies (leukemia and lymphomas), myelodysplastic and -proliferative syndromes, hemoglobinopathies (sickle cell disease and thalassemia), receipt of stem cell transplantation or cell therapy such as CAR T-cell therapy.

***For steroid treatment the exclusion criteria were: receipt of any high-dose (≥ 20 mg of prednisone daily or equivalent) steroid treatment; daily corticosteroids (locally, incl. inhaled steroids, are acceptable) within 2 weeks of study entry; or repeated use of any high dose of corticosteroids (a dose of > 30 mg of prednisone or equivalent per day for multiple days) in the recent past

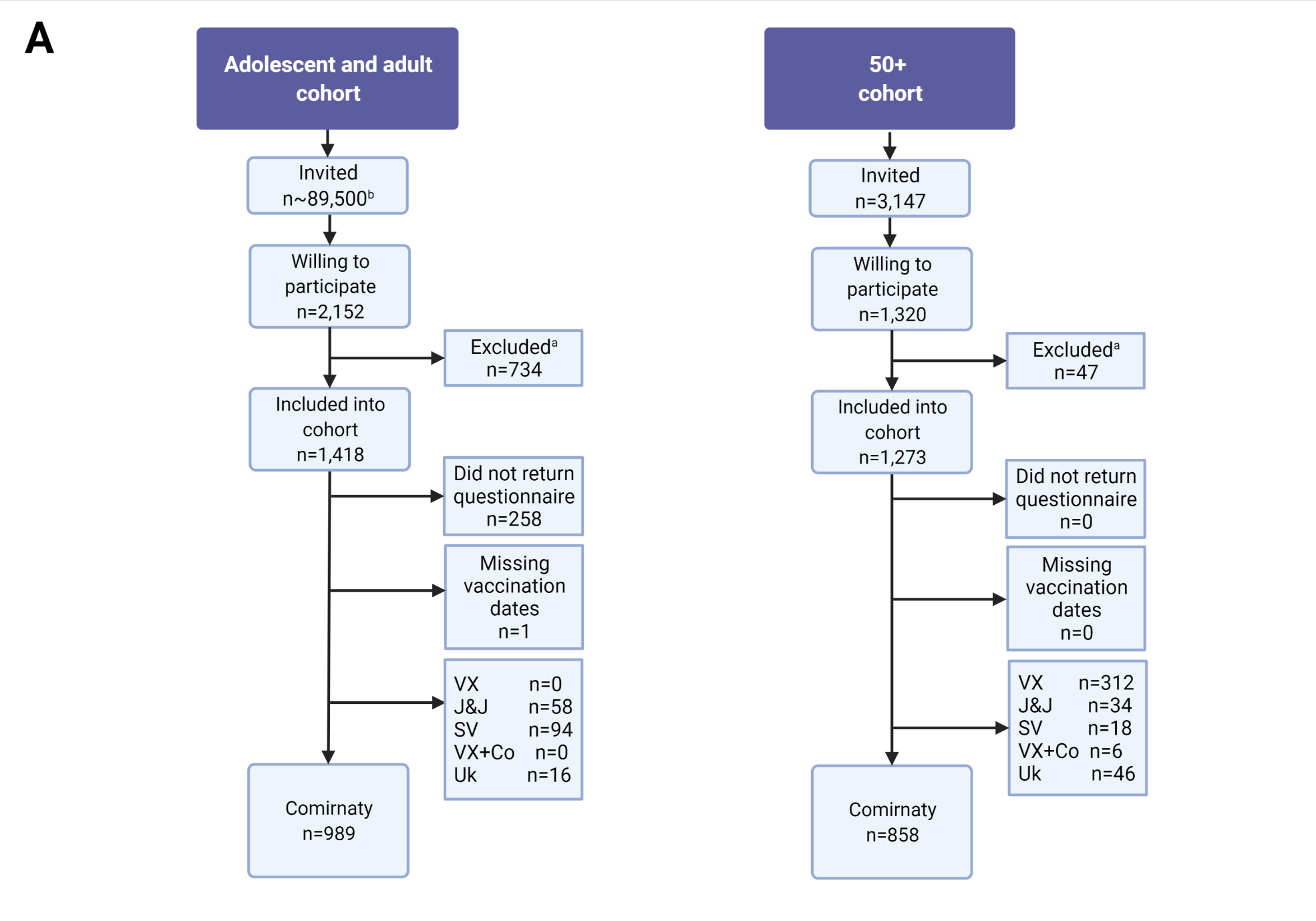

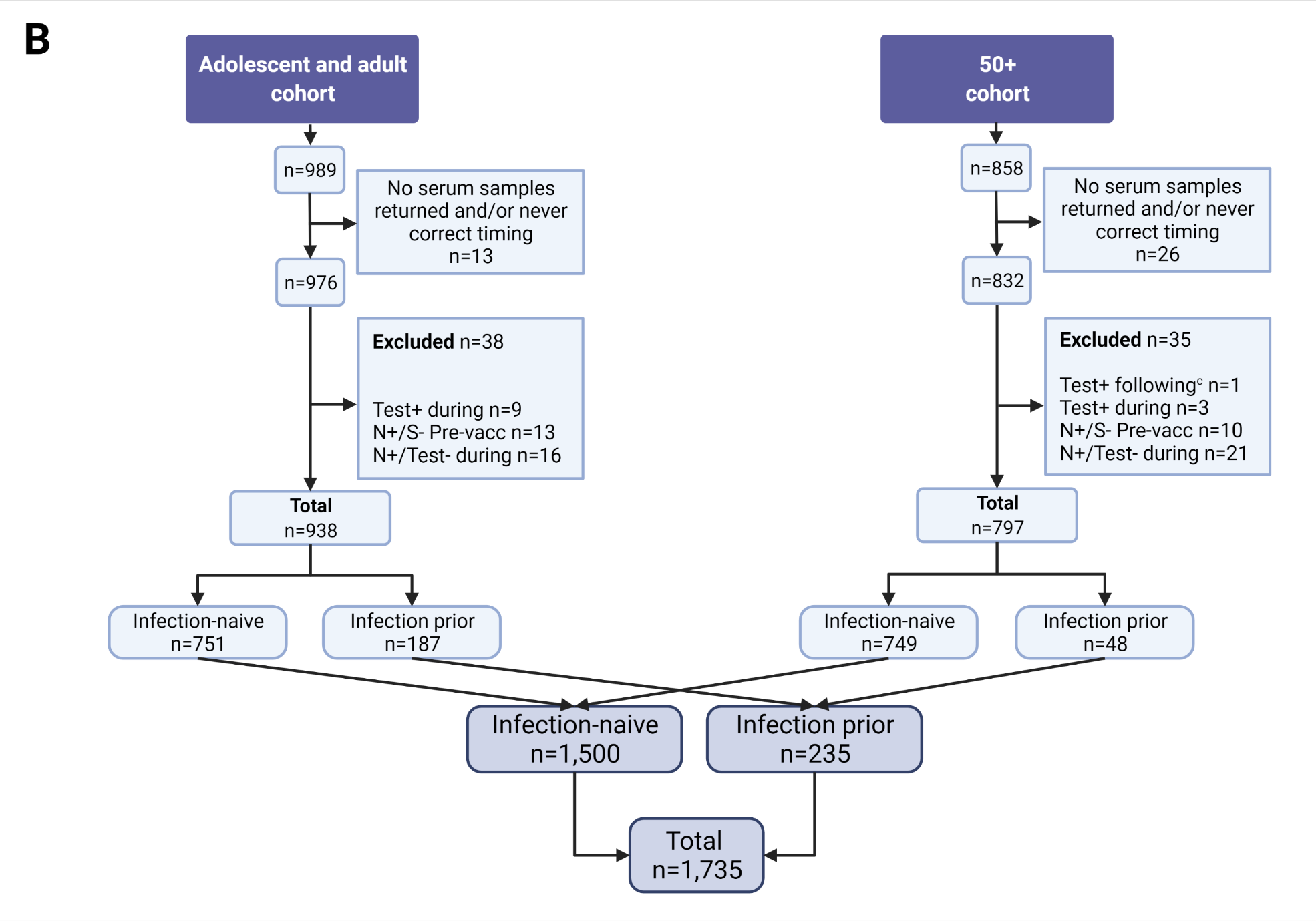

**Supplementary Figure 1: Flowchart of participants included in the study.** In (A) an overview is shown of the number of participants included into each cohort and who received Comirnaty vaccination. In (B) the number of participants without serum samples available and/or never returning a sample in the correct time window are shown as well as exclusions based on Nucleoprotein seroconversion or a positive SARS-CoV-2 test during the vaccination schedule, or those who were only seropositive to Nucleoprotein and not Spike S1 prior to vaccination as their infection-status was considered inconclusive. Participants were categorized into infection-naïve or those with a SARS-CoV-2 infection prior to vaccination (i.e., positive SARS-CoV-2 test and/or Spike S1 seropositive prior to vaccination). ^a^Exclusion based on no further replies by participant after notification of willingness to participate, did not complete informed consent or – for the 12-17 and 18-60 cohorts only – based on exclusion criteria listed in Supplementary Table 1. ^b^In addition to a total of 40,000 persons who were invited into the study by random sampling from the National Population Register (Basis Registratie Personen; BRP), approximately 9,500 participants who participated in prior studies by the Dutch National Institute of Public Health and the Environment (RIVM) and who consented to be contacted for additional research were invited as well as an unknown number of spontaneous enrollments from interested citizens. ^c^For the 50+ cohort, participants with a positive SARS-CoV-2 test following vaccination were excluded as for this population measurements were analyzed up to three months following the second vaccination dose. VX: Vaxzevria; J&J: Janssen; SV: Spikevax; VX+Co: first dose Vaxzevria and second dose Comirnaty; Uk: unknown; Test+ during: positive SARS-CoV-2 test during primary vaccination schedule (antigen or PCR performed by local health authorities); N+/S-: seropositive to Nucleoprotein but seronegative to Spike S1 prior to vaccination; N+/Test-: seropositive to Nucleoprotein but no positive SARS-CoV-2 test reported during vaccination schedule; Test+ following: positive SARS-CoV-2 test following primary vaccination schedule (antigen or PCR performed by local health authorities).

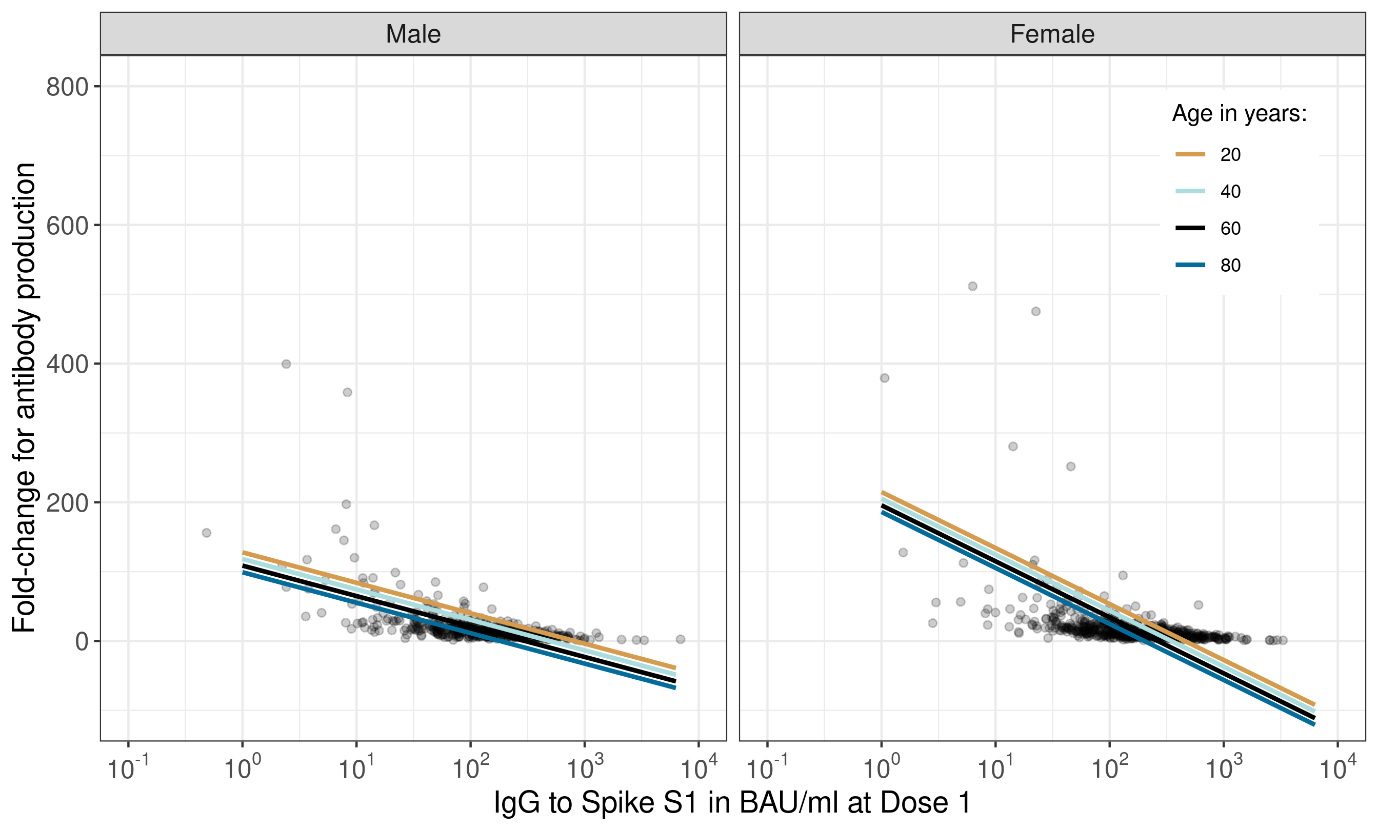

**Supplementary Figure 2: The fold-change in Spike S1-specific IgG to represent antibody production between one month after the first and one month after the second dose of BNT162b2 by S1 IgG concentration at one month after the first dose in infection-naïve participants** **across all ages.** Fitted lines represent the linear association between fold-change and S1 IgG concentration from linear regression for four ages (see Supplementary Table 5), while dots represent individual measurements. IgG concentration measurements were expressed in international binding antibody units (BAU) using the 20/136 NIBSC standard. Results are shown separately for males and females. Participants were only included if they had measurements available at both timepoints (n=1,089).

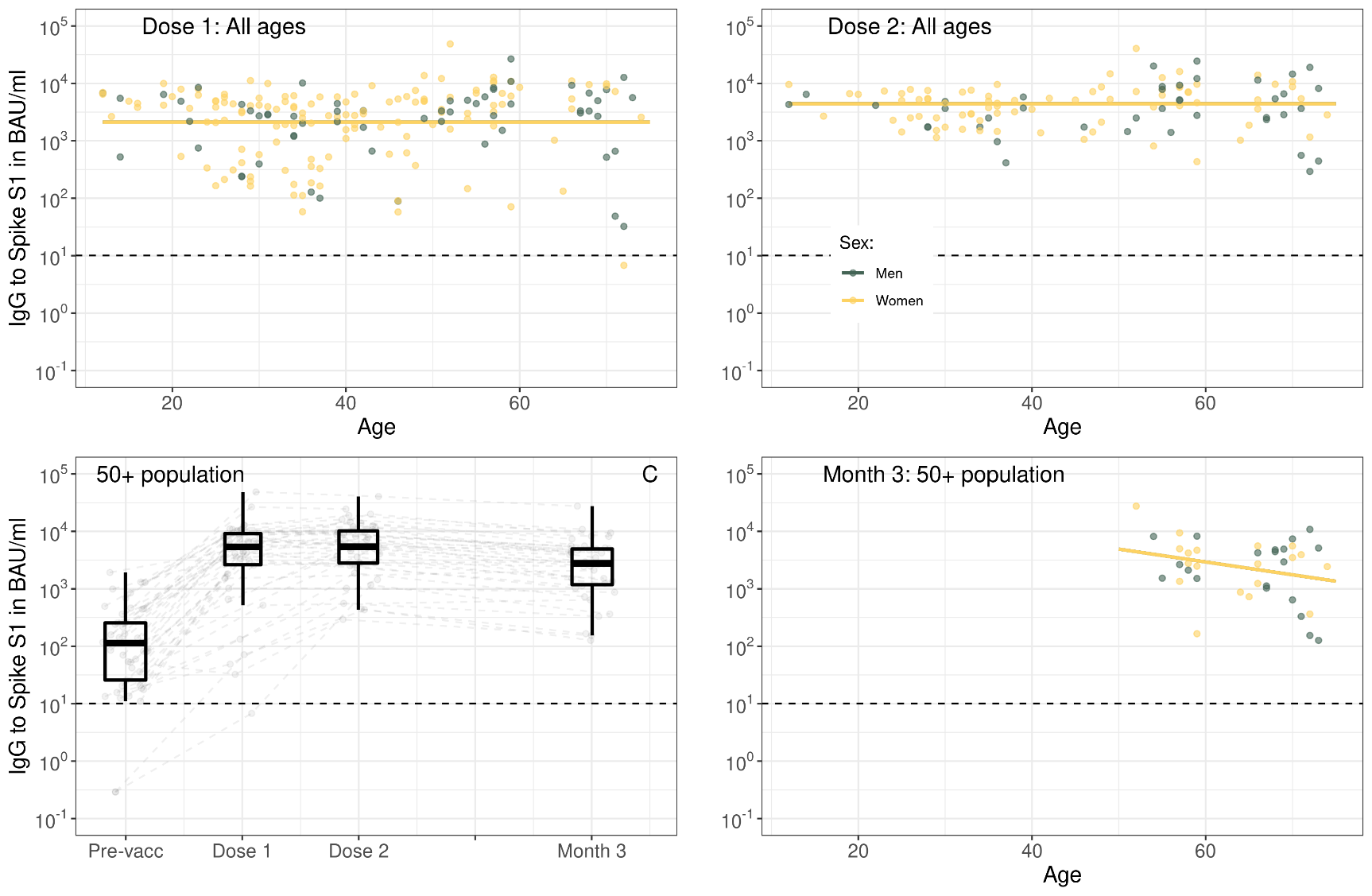

**Supplementary Figure 3: Spike S1-specific IgG by age in years per timepoint (A, B, D) and kinetics following the primary series of BNT162b2 vaccination in participants** **with a history of SARS-CoV-2 infection up to three months following the second vaccination dose (C).** In (A, B, D) fitted lines represent the linear association between IgG concentration and age from linear mixed effects regression results (see Supplementary Table 3 and Supplementary Table 4), while dots represent individual measurements. Results are shown separately for men (green) and women (yellow). In (C) boxplots show results for all participants at each timepoint, while dots and dashed grey lines show measurements and their trajectory between timepoints per participant. In (A-D) IgG concentration measurements were expressed in international binding antibody units (BAU) using the 20/136 NIBSC standard and were taken prior to vaccination (Pre-vacc), one month following the first (Dose 1), one month following the second (Dose 2) or three months following the second vaccination dose (Month 3). In (A-B) results are shown for a total of 204 unique participants across all ages with S1 IgG measurements available at Dose 1 and/or Dose 2, while in (C) results are shown for 48 unique participants in the 50+ cohort and in (D) results are shown for 46 unique participants in the 50+ cohort with measurements available at Dose 2 and/or Month 3. The horizontal dashed line represents the threshold for seropositivity to Spike S1. The horizontal dashed line represents the threshold for seropositivity to Spike S1. IgG: immunoglobulin G; BAU: binding antibody units.

**Supplementary Table 1: General characteristics of the study population one month following one and two doses of BNT162b2 by age category and SARS-CoV-2 infection status.**

|  | **12-19** | **20-29** | **30-39** | **40-49** | **50-59** | **60-69** | **70-79** | **80-93** | **All ages** |
| --- | --- | --- | --- | --- | --- | --- | --- | --- | --- |
| N | 144 | 222 | 306 | 177 | 255 | 294 | 259 | 78 | 1,735 |
| **Infection-naïve** |  |  |  |  |  |  |  |  |  |
| **N, %** | 127, 88% | 174, 78% | 249, 81% | 140, 79% | 207, 81% | 280, 95% | 245, 95% | 78, 100% | 1,500, 86% |
| **Sex*, %** |  |  |  |  |  |  |  |  |  |
| - Male | 47, 37% | 55, 32% | 90, 36% | 42, 30% | 92, 44% | 120, 43% | 119, 49% | 48, 62% | 615, 41% |
| - Female | 78, 61% | 118, 68% | 158, 64% | 97, 69% | 115, 56% | 160, 57% | 126, 51% | 30, 38% | 877, 59% |
| - Other | 2, 2% | 1, 1% | 1, 0% | 1, 1% | 0, 0% | 0, 0% | 0, 0% | 0, 0% | 5, 0% |
| **Vaccination interval in days** |  |  |  |  |  |  |  |  |  |
| - N with two doses | 126, 99% | 173, 99% | 248, 100% | 139, 99% | 207, 100% | 280, 100% | 245, 100% | 78, 100% | 1,496, 100% |
| - Median | 35 | 35 | 35 | 35 | 35 | 35 | 35 | 35 | 35 |
| - IQR | 23 – 35 | 35 – 35 | 35 – 36 | 35 – 36 | 35 – 35 | 35 – 35 | 35 – 35 | 35 – 35 | 35 – 35 |
| **History of SARS-CoV-2 infection** |  |  |  |  |  |  |  |  |  |
| **N, %** | 17, 12% | 48, 22% | 57, 19% | 37, 21% | 48, 19% | 14, 5% | 14, 5% | 0, 0% | 235, 14% |
| **Sex*, %** |  |  |  |  |  |  |  |  |  |
| - Male | 5, 29% | 12, 25% | 15, 26% | 7, 19% | 21, 44% | 7, 50% | 8, 57% | N/A | 75, 32% |
| - Female | 12, 71% | 36, 75% | 42, 74% | 30, 81% | 27, 56% | 7, 50% | 6, 43% | N/A | 160, 68% |
| - Other | 0, 0% | 0, 0% | 0, 0% | 0, 0% | 0, 0% | 0, 0% | 0, 0% | N/A | 0, 0% |
| **Vaccination interval** |  |  |  |  |  |  |  |  |  |
| - N with two doses | 9, 53% | 30, 63% | 37, 65% | 18, 49% | 39, 82% | 13, 92% | 14, 100% | N/A | 160, 68% |
| - Median | 28 | 35 | 35 | 35 | 35 | 35 | 35 | N/A | 35 |
| - IQR | 22 – 35 | 35 – 38 | 35 – 38 | 35 – 40 | 35 – 36 | 35 – 36 | 35 – 35 | N/A | 35 – 37 |

*In the adolescents and adult cohort study (ages 12 to 60), participants could indicate male, female or other. In the 50+ cohort, participants could only indicate male or female.

**Supplementary Table 2: Spike S1 seropositivity and IgG concentrations one month following one and two doses of BNT162b2 by age category and SARS-CoV-2 infection status; and at three months following the second dose for the 50+ cohort.**

|  | **12-19** | **20-29** | **30-39** | **40-49** | **50-59** | **60-69** | **70-79** | **80-93** | **All ages** |
| --- | --- | --- | --- | --- | --- | --- | --- | --- | --- |
| **Infection-naïve** |  |  |  |  |  |  |  |  |  |
| **Seropositivity IgG to S1 (n/N)** |  |  |  |  |  |  |  |  |  |
| - Dose 1 | 100%  (56/56) | 100% (148/148) | 100% (207/207) | 99% (124/125) | 99% (176/178) | 97% (238/245) | 94% (206/219) | 86%  (49/57) | 98%  (1,204/1,235) |
| - Dose 2 | 100% (107/107) | 100% (141/141) | 100% (190/190) | 100% (116/116) | 100% (182/182) | 100% (265/265) | 100% (234/234) | 100%  (73/73) | 100% (1,308/1,308) |
| - Month 3 | N/A | N/A | N/A | N/A | 100% (116/116) | 100% (249/249) | 100% (222/222) | 100%  (68/68) | 100%  (655/655) |
| **Median IgG to S1 (IQR)** |  |  |  |  |  |  |  |  |  |
| - Dose 1 | 538  (284-805) | 315  (221-594) | 204  (112-301) | 154  (79-265) | 127  (63-231) | 109  (60-215) | 96  (42-176) | 45  (16-113) | 146  (72-290) |
| - Dose 2 | 4,218  (2,707-6,174) | 2,972  (2,074-4,411) | 2,177  (1,490-3,509) | 1,673  (1,056-2,562) | 1,765  (1,010-2,813) | 1,435  (815-2,421) | 1,238  (791-2,080) | 672  (366-1,304) | 1,842  (1,019-3,116) |
| - Month 3 | N/A | N/A | N/A | N/A | 486  (312-898) | 456  (234-737) | 446  (263-750) | 210  (127-519) | 440  (239-736) |
| **History of SARS-CoV-2 infection** |  |  |  |  |  |  |  |  |  |
| **Seropositivity IgG to S1 (n/N)** |  |  |  |  |  |  |  |  |  |
| - Pre-vacc | 87%  (13/15) | 85%  (41/48) | 86%  (49/57) | 91%  (32/35) | 89%  (41/46) | 93%  (13/14) | 92%  (11/12) | N/A | 100%  (200/227) |
| - Dose 1 | 100%  (12/12) | 100%  (37/37) | 100%  (48/48) | 100%  (33/33) | 100%  (38/38) | 100%  (14/14) | 92%  (11/12) | N/A | 100%  (193/194) |
| - Dose 2 | 100%  (6/6) | 100%  (18/18) | 100%  (28/28) | 100%  (12/12) | 100%  (29/29) | 100%  (12/12) | 100%  (12/12) | N/A | 100%  (117/117) |
| - Month 3 | N/A | N/A | N/A | N/A | 100%  (15/15) | 100%  (12/12) | 100%  (12/12) | N/A | 100%  (39/39) |
| **Median IgG to S1 (IQR)** |  |  |  |  |  |  |  |  |  |
| - Pre-vacc | 153  (29-271) | 63  (34-105) | 55  (28-104) | 83  (37-139) | 82  (28-235) | 128  (19-204) | 62  (33-199) | N/A | 73  (29-147) |
| - Dose 1 | 5,192  (4,085-6,561) | 3,286  (416-4,884) | 2,325  (457-3,941) | 2,760  (1,190-4,862) | 4,405  (2,814-7,810) | 4,296  (3,112-8,090) | 4,111  (401-8,085) | N/A | 3,293  (1,191-5,751) |
| - Dose 2 | 6,446  (4,832-6,583) | 3,404  (1,887-5,424) | 3,425  (2,123-4,857) | 4,336  (1,657-5,925) | 7,119  (3,886-9,319) | 4,375  (2,486-7,336) | 4,531  (1,011-9,263) | N/A | 4,535  (2,341-7,205) |
| - Month 3 | N/A | N/A | N/A | N/A | 2,774  (1,820-6,574) | 2,827  (1,100-4,521) | 2,967  (355-5,244) | N/A | 2,774  (1,182-4,948) |

**Supplementary Table 3: Linear regression results for fold-change in Spike S1 IgG concentrations at one month after the first and one month after the second dose in infection-naïve participants** **across all ages.** The dependent variable was the fold-change for antibody production which was calculated by dividing the IgG S1 concentration at Dose 2 by the IgG S1 concentration at Dose 1. Participants were only included if they had measurements available at both time points (n=1,089).

|  | **Coefficient** | **95% CI** | **p-value** |
| --- | --- | --- | --- |
| **IgG S1 concentration at Dose 1** | -43.914 | -60.551,-27.276 | <0.001 |
| **Age in years** | -0.479 | -0.794,-0.164 | 0.003 |
| **Sex** |  |  |  |
| - Male | Ref. |  |  |
| - Female | 86.976 | 40.710,133.241 | <0.001 |
| **IgG S1 concentration at Dose 1*Sex** |  |  |  |
| - IgG S1 concentration at Dose 2*Male | Ref. |  |  |
| - IgG S1 concentration at Dose 2*Female | -36.866 | -58.229,-15.503 | 0.001 |

**Supplementary Table 4: Linear mixed effects regression results for Spike S1 IgG concentrations up to one month following two doses of BNT162b2 for participants** **with a SARS-CoV-2 infection history.** Results are shown for total of 204 unique participants across all ages with S1 IgG measurements available at Dose 1 and/or Dose 2.

|  | **Coefficient** | **95% CI** | **p-value** |
| --- | --- | --- | --- |
| **Timepoint** |  |  |  |
| - Pre-vacc | -1.614 | -1.726,-1.501 | <0.001 |
| - Dose 1 | Ref. |  |  |
| - Dose 2 | 0.260 | 0.122,0.398 | <0.001 |

**Supplementary Table 5: Linear mixed effects regression results for Spike S1 IgG concentrations one and three months following two doses of BNT162b2 by prior SARS-CoV-2 infection status in the 50+ cohort.** Results are shown for total of 725 unique infection-naïve and 46 unique participants with a SARS-CoV-2 infection history in the 50+ cohort with measurements available at Dose 2 and/or Month 3.

|  | **Infection-naïve** |  |  | **History of SARS-CoV-2 infection** |  |  |
| --- | --- | --- | --- | --- | --- | --- |
|  | **Coefficient** | **95% CI** | **p-value** | **Coefficient** | **95% CI** | **p-value** |
| **Timepoint** |  |  |  |  |  |  |
| - Dose 2 | Ref. |  |  | Ref. |  |  |
| - Month 3 | -0.792 | -0.950,-0.635 | <0.001 | -0.299 | -0.350,-0.248 | <0.001 |
| **Age in years** | -0.012 | -0.016,-0.009 | <0.001 |  |  |  |
| **Sex** |  |  |  |  |  |  |
| - Male | Ref. |  |  |  |  |  |
| - Female | 0.102 | 0.053,0.151 | <0.001 |  |  |  |
| **Age*Timepoint** |  |  |  |  |  |  |
| - Age*Dose 2 | Ref. |  |  |  |  |  |
| - Age*Month 3 | 0.004 | 0.002,0.007 | <0.001 |  |  |  |

**Supplementary Table 6: Linear regression results for fold-change in Spike S1 IgG concentrations at one month and three months following two doses of BNT162b2 in infection-naïve participants** **from the 50+ cohort.** The dependent variable was the fold-change for antibody loss which was calculated by dividing the IgG S1 concentration at Month 3 by the IgG S1 concentration at Dose 2. Participants were only included if they had measurements available at both time points (n=633).

|  | **Coefficient** | **95% CI** | **p-value** |
| --- | --- | --- | --- |
| **IgG S1 concentration at Dose 1** | -0.099 | -0.168,-0.031 | 0.004 |
| **Age in years** | 0.003 | 0.001,0.005 | 0.007 |
| **Sex** |  |  |  |
| - Male | Ref. |  |  |
| - Female | 0.322 | 0.041,0.603 | 0.025 |
| **IgG S1 concentration at Dose 1*Sex** |  |  |  |
| - IgG S1 concentration at Dose 2*Male | Ref. |  |  |
| - IgG S1 concentration at Dose 2*Female | -0.091 | -0.180,-0.001 | 0.048 |
